# Effectiveness of BNT162b2 and mRNA-1273 Second Doses and Boosters for SARS-CoV-2 infection and SARS-CoV-2 Related Hospitalizations: A Statewide Report from the Minnesota Electronic Health Record Consortium

**DOI:** 10.1101/2021.12.23.21267853

**Authors:** Paul E Drawz, Malini DeSilva, Peter Bodurtha, Gabriela Vazquez Benitez, Anne Murray, Alanna M Chamberlain, R Adams Dudley, Stephen Waring, Anupam B Kharbanda, Daniel Murphy, Miriam Halstead Muscoplat, Victor Melendez, Karen L Margolis, Lynn McFarling, Roxana Lupu, Tyler N.A. Winkelman, Steve Johnson

## Abstract

Using vaccine data combined with electronic health records, we report that mRNA boosters provide greater protection than a two-dose regimen against SARS-CoV-2 infection and related hospitalizations. The benefit of a booster was more evident in the elderly and those with comorbidities. These results support the case for COVID-19 boosters.

---

COVID-19 vaccines are effective at reducing SARS-CoV-2 infections and related hospitalizations but there is concern for waning immunity.^1^ COVID-19 vaccine booster doses increase antibodies and cellular responses.^2^ However, data on the effectiveness of boosters are limited.^3-5^

We report on vaccine effectiveness (VE) in individuals who received a second dose of a BNT162b2 (Pfizer-BioNTech) or mRNA-1273 (Moderna) COVID-19 vaccine and in those who received a booster dose of either vaccine. We used statewide COVID-19 vaccination data from the Minnesota Immunization Information Connection (MIIC) linked via a privacy-preserving record linkage process with distributed electronic health record (EHR) data from the 11 largest health systems in Minnesota.^6^ Health systems reported data aggregated by categories defined by race/ethnicity, age, sex, and comorbidities. The Institutional Review Board (IRB) of Mayo Clinic gave ethical approval for this work. The IRB of CentraCare, the University of Minnesota, Hennepin Healthcare, Children’s Minnesota, Allina Health, the Minneapolis VA, Sanford Health, HealthPartners, and Essentia Health waived ethical approval of this work as it qualifies as public health surveillance. Investigators and the legal office at NorthMemorial deemed this work to be public health surveillance.^7,8^ Participating sites care for 50% of SARS-CoV-2 positive cases across Minnesota and the health systems represent approximately 75% of hospital admissions. Individuals with no matching MIIC vaccination record were considered unvaccinated. Individuals were considered fully vaccinated after receipt of a second dose of a Pfizer or Moderna vaccine or a single dose of an Ad26.COV.2.S (Janssen) vaccine. The next COVID-19 dose after an individual was fully vaccinated was defined as a booster dose.

VE for Pfizer and Moderna vaccines was estimated among those age 19 years and older using a test-negative design for infections and incident rate ratio (IRR) for hospitalizations among individuals greater than 26 weeks after being fully vaccinated (the time point at which individuals are eligible for a booster) and those who had received a booster. For the test-negative design, we utilized all available SARS-CoV-2 PCR test results. For IRR, the outcome was a hospital admission the same week or within three weeks after a positive SARS-CoV-2 PCR test. Unvaccinated individuals were the comparison group for all analyses. Analyses were unadjusted and limited to demographic groups and those with high risk conditions with at least 6 events and more than 25,000 person-weeks at risk. We evaluated Morbidity and Mortality Weekly Report (MMWR) weeks 35 through 47 (August 29 through November 27) to capture the period when booster doses were recommended.

Of 4,547,945 patients from participating health systems, 1,732,112 were fully vaccinated with Pfizer and 1,066,645 were fully vaccinated with Moderna (Supplemental Table 1). A Pfizer booster was administered to 609,153 individuals; a Moderna booster was administered to 395,634 individuals. VE using a test-negative design for individuals greater than 26 weeks from a second dose was 45% (95% CI 44 to 47) for Pfizer and 65% (95% CI 65 to 66) for Moderna (Figure 1a, Supplemental Table 2). For individuals who had received a booster dose, VE was 88% (95% CI 87 to 88) for Pfizer and 91% (95% CI 90 to 92) for Moderna. VE for SARS-CoV-2 related hospitalizations for individuals greater than 26 weeks from a second dose was 67% (95% CI 65 to 69) for Pfizer and 73% (95% CI 71 to 75) for Moderna (Figure 1b, Supplemental Table 3). VE for SARS-CoV-2 related hospitalizations for boosters was 88% (95% CI 86 to 90) for Pfizer and 86% (95% CI 82 to 89) for Moderna. The benefit of a booster was more evident in the elderly and those with comorbidities.

**Figure 1.**
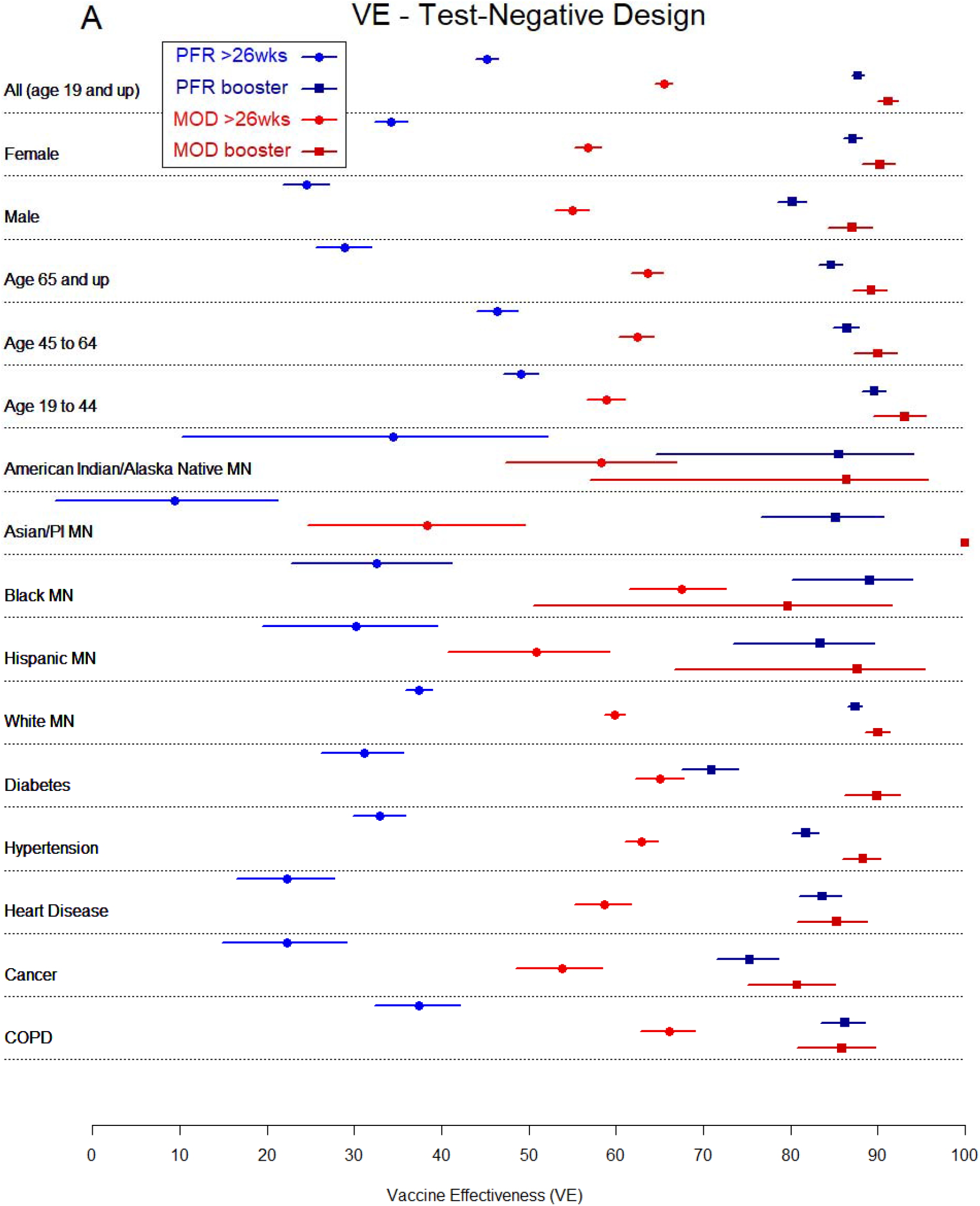
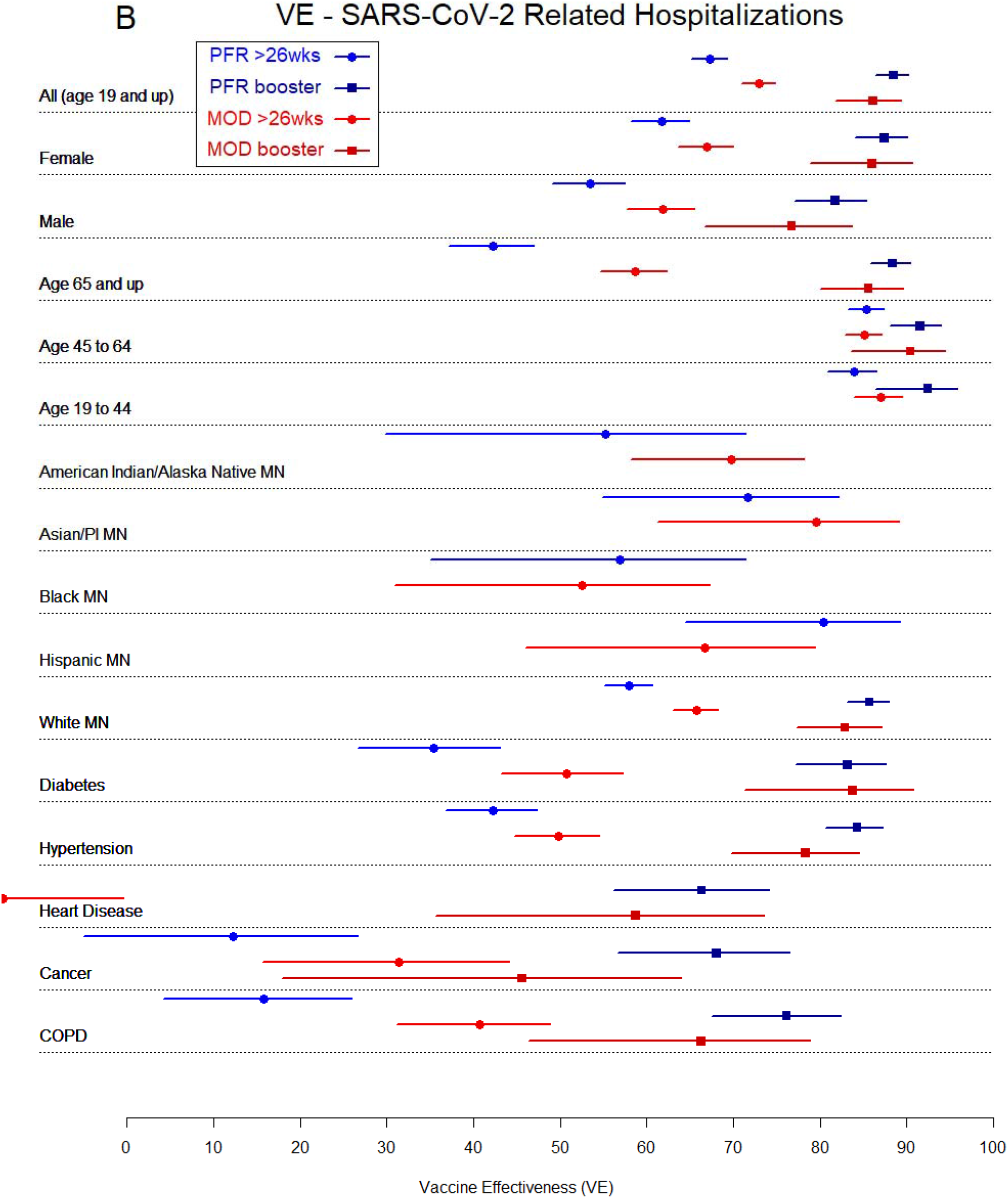
Vaccine effectiveness overall and by subgroups as assessed by SARS-CoV-2 test positivity (test-negative design (100 * (1 – OR)), A) and SARS-CoV-2 related hospitalizations (100 * (1 – IRR), B). August 29 to November 27, 2021.

Using statewide vaccine data combined with EHR data in Minnesota, we report that mRNA booster doses appear to provide greater protection against medically attended SARS-CoV-2 infection and SARS-CoV-2 related hospitalizations. Our results are consistent with prior studies from Israel that only reported on the VE of Pfizer booster doses.^3-5^

There are some limitations to consider. First, it is possible that our definition of a SARS-CoV-2 related hospitalization may include admissions unrelated to the infection. Second, while results are consistent across subgroups, because health systems aggregated results by categories of single variables, we cannot provide multi-variable adjusted analyses. Finally, while the Minnesota EHR Consortium data includes most of the large health systems in Minnesota, the analyses do not include complete statewide data.

In conclusion, we demonstrate that individuals who have received a booster have a greater degree of protection against SARS-CoV-2 infection and related hospitalizations compared to individuals greater than 26 weeks from their final dose of either a Moderna or Pfizer vaccine. These results can inform the distribution of COVID-19 booster doses.

## Data Availability

Data produced in the present study may be requested. All requests must be approved by the Minnesota Electronic Health Record Consortium Steering Committee.

