## Supplemental Table 1 for "Effectiveness of BNT162b2 and mRNA-1273 Second Doses and Boosters for SARS-CoV-2 infection and SARS-CoV-2 Related Hospitalizations: A Statewide Report from the Minnesota Electronic Health Record Consortium"

Supplemental Table 1. Characteristics of individuals who received a second dose of a Pfizer or Moderna vaccine and those who received a booster dose (through November 27, 2021).

| | Received Second Dose | | Booster Dose | |
|---|---|---|---|---|
| Group | Pfizer | Moderna | Pfizer | Moderna |
| Total | 1,732,112 | 1,066,645 | 609,153 | 395,634 |
| Age 0 to 18 | 248,614 | 7,186 | 2,898 | 608 |
| Age 19 to 24 | 108,668 | 69,760 | 11,621 | 8,198 |
| Age 25 to 44 | 453,938 | 301,836 | 113,902 | 69,049 |
| Age 45 to 64 | 501,756 | 362,835 | 179,092 | 124,029 |
| Age 65 to 74 | 223,685 | 196,951 | 156,704 | 122,144 |
| Age 75 and up | 193,561 | 127,184 | 144,688 | 71,479 |
| Age missing | 1,890 | 893 | 248 | 127 |
| Female | 935,923 | 581,345 | 352,002 | 226,975 |
| Male | 796,189 | 485,300 | 257,151 | 168,659 |
| Asian/Pacific Islander | 101,361 | 48,984 | 22,872 | 11,827 |
| Black | 97,753 | 52,980 | 12,550 | 7,749 |
| Hispanic | 72,944 | 37,904 | 10,476 | 5,938 |
| Multiracial | 19,630 | 8,912 | 3,688 | 2,387 |
| American Indian or Alaska Native | 9,494 | 11,407 | 2,036 | 3,756 |
| White | 1,364,534 | 859,030 | 540,456 | 349,267 |
| Other, Unknown, Missing | 66,396 | 47,428 | 17,075 | 14,710 |
| Diabetes | 124,347 | 87,984 | 68,869 | 40,890 |
| Hypertension | 341,705 | 238,458 | 200,346 | 117,881 |
| Heart Disease | 109,689 | 81,479 | 70,803 | 41,715 |
| Cancer | 101,759 | 66,397 | 65,795 | 37,165 |
| COPD | 91,618 | 63,395 | 44,638 | 27,558 |

Supplemental Table 2. Vaccine effectiveness for SARS-CoV-2 infections by manufacturer and booster status

| | | | Vaccinated | | Unvaccinated | | | | |
|---|---|---|---|---|---|---|---|---|---|
| Group | Manufacturer | Status | Tested | Positive | Tested | Positive | VE | VE 95% CI LL | VE 95% CI UL |
| All (age 19 and up) | PFR | Complete >26 | 81486 | 9823 | 209537 | 41956 | 45 | 44 | 47 |
| All (age 19 and up) | PFR | booster | 38291 | 1141 | 209537 | 41956 | 88 | 87 | 88 |
| All (age 19 and up) | MOD | Complete >26 | 73934 | 5879 | 209537 | 41956 | 65 | 64 | 66 |
| All (age 19 and up) | MOD | booster | 9582 | 206 | 209537 | 41956 | 91 | 90 | 92 |
| Female | PFR | Complete >26 | 53162 | 5882 | 213099 | 33912 | 34 | 32 | 36 |
| Female | PFR | booster | 23832 | 566 | 213099 | 33912 | 87 | 86 | 88 |
| Female | MOD | Complete >26 | 45770 | 3458 | 213099 | 33912 | 57 | 55 | 58 |
| Female | MOD | booster | 5640 | 102 | 213099 | 33912 | 90 | 88 | 92 |
| Male | PFR | Complete >26 | 29647 | 4039 | 195677 | 33845 | 25 | 22 | 27 |
| Male | PFR | booster | 14631 | 581 | 195677 | 33845 | 80 | 78 | 82 |
| Male | MOD | Complete >26 | 28335 | 2435 | 195677 | 33845 | 55 | 53 | 57 |
| Male | MOD | booster | 3944 | 104 | 195677 | 33845 | 87 | 84 | 89 |
| Age 65 and up | PFR | Complete >26 | 30027 | 3663 | 33585 | 5488 | 29 | 26 | 32 |
| Age 65 and up | PFR | booster | 19330 | 562 | 33585 | 5488 | 85 | 83 | 86 |
| Age 65 and up | MOD | Complete >26 | 37306 | 2473 | 33585 | 5488 | 64 | 62 | 65 |
| Age 65 and up | MOD | booster | 5837 | 120 | 33585 | 5488 | 89 | 87 | 91 |
| Age 45 to 64 | PFR | Complete >26 | 23898 | 2997 | 56798 | 11993 | 46 | 44 | 49 |
| Age 45 to 64 | PFR | booster | 9919 | 346 | 56798 | 11993 | 86 | 85 | 88 |
| Age 45 to 64 | MOD | Complete >26 | 19785 | 1809 | 56798 | 11993 | 62 | 60 | 64 |
| Age 45 to 64 | MOD | booster | 2470 | 64 | 56798 | 11993 | 90 | 87 | 92 |
| Age 19 to 44 | PFR | Complete >26 | 27553 | 3163 | 116206 | 23603 | 49 | 47 | 51 |
| Age 19 to 44 | PFR | booster | 9036 | 233 | 116206 | 23603 | 90 | 88 | 91 |
| Age 19 to 44 | MOD | Complete >26 | 16841 | 1597 | 116206 | 23603 | 59 | 57 | 61 |
| Age 19 to 44 | MOD | booster | 1272 | 22 | 116206 | 23603 | 93 | 89 | 95 |
| American Indian/Alaska Native MN | PFR | Complete >26 | 346 | 47 | 5110 | 989 | 35 | 10 | 52 |
| American Indian/Alaska Native MN | PFR | booster | 149 | <11 | 5110 | 989 | 85 | 65 | 95 |
| American Indian/Alaska Native MN | MOD | Complete >26 | 934 | 85 | 5110 | 989 | 58 | 47 | 67 |
| American Indian/Alaska Native MN | MOD | booster | 95 | <11 | 5110 | 989 | 85 | 55 | 95 |
| Asian/PI MN | PFR | Complete >26 | 2502 | 257 | 12328 | 1383 | 9 | -4 | 21 |
| Asian/PI MN | PFR | booster | 1036 | 19 | 12328 | 1383 | 85 | 77 | 91 |
| Asian/PI MN | MOD | Complete >26 | 1550 | 112 | 12328 | 1383 | 38 | 25 | 50 |
| Asian/PI MN | MOD | booster | 165 | <11 | 12328 | 1383 | 100 | | 100 |
| Black MN | PFR | Complete >26 | 2569 | 239 | 44373 | 5859 | 33 | 23 | 41 |
| Black MN | PFR | booster | 675 | 11 | 44373 | 5859 | 89 | 80 | 94 |
| Black MN | MOD | Complete >26 | 3021 | 142 | 44373 | 5859 | 68 | 62 | 73 |
| Black MN | MOD | booster | 167 | <11 | 44373 | 5859 | 80 | 50 | 90 |
| Hispanic MN | PFR | Complete >26 | 1929 | 222 | 26643 | 4186 | 30 | 19 | 40 |
| Hispanic MN | PFR | booster | 600 | 18 | 26643 | 4186 | 83 | 73 | 90 |
| Hispanic MN | MOD | Complete >26 | 1478 | 124 | 26643 | 4186 | 51 | 41 | 59 |
| Hispanic MN | MOD | booster | 178 | <11 | 26643 | 4186 | 90 | 65 | 95 |
| White MN | PFR | Complete >26 | 73090 | 8927 | 275652 | 50170 | 37 | 36 | 39 |
| White MN | PFR | booster | 35060 | 955 | 275652 | 50170 | 87 | 87 | 88 |
| White MN | MOD | Complete >26 | 64217 | 5264 | 275652 | 50170 | 60 | 59 | 61 |
| White MN | MOD | booster | 8691 | 188 | 275652 | 50170 | 90 | 89 | 91 |
| Diabetes | PFR | Complete >26 | 10209 | 1398 | 16622 | 3112 | 31 | 26 | 36 |
| Diabetes | PFR | booster | 5850 | 367 | 16622 | 3112 | 71 | 67 | 74 |
| Diabetes | MOD | Complete >26 | 11522 | 858 | 16622 | 3112 | 65 | 62 | 68 |
| Diabetes | MOD | booster | 1805 | 41 | 16622 | 3112 | 90 | 86 | 93 |
| Hypertension | PFR | Complete >26 | 26113 | 3412 | 38510 | 7051 | 33 | 30 | 36 |
| Hypertension | PFR | booster | 15478 | 607 | 38510 | 7051 | 82 | 80 | 83 |
| Hypertension | MOD | Complete >26 | 27224 | 2087 | 38510 | 7051 | 63 | 61 | 65 |
| Hypertension | MOD | booster | 4475 | 114 | 38510 | 7051 | 88 | 86 | 90 |
| Heart Disease | PFR | Complete >26 | 12990 | 1418 | 15363 | 2093 | 22 | 17 | 28 |
| Heart Disease | PFR | booster | 8065 | 203 | 15363 | 2093 | 84 | 81 | 86 |
| Heart Disease | MOD | Complete >26 | 16070 | 984 | 15363 | 2093 | 59 | 55 | 62 |
| Heart Disease | MOD | booster | 2434 | 55 | 15363 | 2093 | 85 | 81 | 89 |
| Cancer | PFR | Complete >26 | 7870 | 842 | 10088 | 1348 | 22 | 15 | 29 |
| Cancer | PFR | booster | 6520 | 239 | 10088 | 1348 | 75 | 72 | 79 |
| Cancer | MOD | Complete >26 | 7484 | 498 | 10088 | 1348 | 54 | 49 | 58 |
| Cancer | MOD | booster | 2153 | 62 | 10088 | 1348 | 81 | 75 | 85 |
| COPD | PFR | Complete >26 | 7664 | 883 | 20530 | 3537 | 37 | 32 | 42 |
| COPD | PFR | booster | 4279 | 119 | 20530 | 3537 | 86 | 83 | 89 |
| COPD | MOD | Complete >26 | 8633 | 569 | 20530 | 3537 | 66 | 63 | 69 |
| COPD | MOD | booster | 1437 | 41 | 20530 | 3537 | 86 | 81 | 90 |

Note: MOD, Moderna; PFR, Pfizer; VE, vaccine effectiveness. Complete >26 indicates individuals more than 26 weeks since a second dose. Booster indicates individuals at least two weeks after a booster dose of Pfizer or Moderna.

Supplemental Table 3. Vaccine effectiveness for SARS-CoV-2 related hospitalizations by manufacturer and booster status

| | | | Vaccinated | | Unvaccinated | | | | |
|---|---|---|---|---|---|---|---|---|---|
| Group | Manufacturer | Status | Person weeks at risk | Hospitalizations | Person weeks at risk | Hospitalizations | VE | VE 95% CI LL | VE 95% CI UL |
| All (age 19 and up) | PFR | Complete >26 | 5955126 | 1093 | 12199893 | 6850 | 67 | 65 | 69 |
| All (age 19 and up) | PFR | Booster | 2289954 | 148 | 12199893 | 6850 | 88 | 86 | 90 |
| All (age 19 and up) | MOD | Complete >26 | 5745746 | 872 | 12199893 | 6850 | 73 | 71 | 75 |
| All (age 19 and up) | MOD | Booster | 680944 | 53 | 12199893 | 6850 | 86 | 82 | 89 |
| Female | PFR | Complete >26 | 3581126 | 573 | 9766332 | 4083 | 62 | 58 | 65 |
| Female | PFR | Booster | 1351212 | 71 | 9766332 | 4083 | 87 | 84 | 90 |
| Female | MOD | Complete >26 | 3377278 | 466 | 9766332 | 4083 | 67 | 64 | 70 |
| Female | MOD | Booster | 392602 | 23 | 9766332 | 4083 | 86 | 79 | 91 |
| Male | PFR | Complete >26 | 2526910 | 525 | 9578393 | 4280 | 54 | 49 | 58 |
| Male | PFR | Booster | 943663 | 77 | 9578393 | 4280 | 82 | 77 | 85 |
| Male | MOD | Complete >26 | 2387611 | 407 | 9578393 | 4280 | 62 | 58 | 66 |
| Male | MOD | Booster | 288447 | 30 | 9578393 | 4280 | 77 | 67 | 84 |
| Age 65 and up | PFR | Complete >26 | 2054120 | 754 | 3038755 | 1933 | 42 | 37 | 47 |
| Age 65 and up | PFR | Booster | 1403149 | 104 | 3038755 | 1933 | 88 | 86 | 90 |
| Age 65 and up | MOD | Complete >26 | 2194729 | 577 | 3038755 | 1933 | 59 | 55 | 62 |
| Age 65 and up | MOD | Booster | 403509 | 37 | 3038755 | 1933 | 86 | 80 | 90 |
| Age 45 to 64 | PFR | Complete >26 | 1914081 | 208 | 3175481 | 2367 | 85 | 83 | 87 |
| Age 45 to 64 | PFR | Booster | 525594 | 33 | 3175481 | 2367 | 92 | 88 | 94 |
| Age 45 to 64 | MOD | Complete >26 | 1840126 | 204 | 3175481 | 2367 | 85 | 83 | 87 |
| Age 45 to 64 | MOD | Booster | 182701 | 13 | 3175481 | 2367 | 90 | 84 | 94 |
| Age 19 to 44 | PFR | Complete >26 | 2007128 | 131 | 5926861 | 2417 | 84 | 81 | 87 |
| Age 19 to 44 | PFR | Booster | 358255 | 11 | 5926861 | 2417 | 92 | 86 | 96 |
| Age 19 to 44 | MOD | Complete >26 | 1715577 | 91 | 5926861 | 2417 | 87 | 84 | 89 |
| American Indian/Alaska Native MN | PFR | Complete >26 | 27892 | 20 | 246492 | 395 | 55 | 30 | 71 |
| American Indian/Alaska Native MN | MOD | Complete >26 | 82577 | 40 | 246492 | 395 | 70 | 58 | 78 |
| Asian/PI MN | PFR | Complete >26 | 289451 | 20 | 673203 | 164 | 72 | 55 | 82 |
| Asian/PI MN | MOD | Complete >26 | 200533 | <11 | 673203 | 164 | 80 | 60 | 90 |
| Black MN | PFR | Complete >26 | 229489 | 24 | 2113897 | 513 | 57 | 35 | 71 |
| Black MN | MOD | Complete >26 | 251666 | 29 | 2113897 | 513 | 53 | 31 | 67 |
| Hispanic MN | PFR | Complete >26 | 167061 | 11 | 1315085 | 443 | 80 | 64 | 89 |
| Hispanic MN | MOD | Complete >26 | 151699 | 17 | 1315085 | 443 | 67 | 46 | 80 |
| White MN | PFR | Complete >26 | 5156692 | 1013 | 13825144 | 6465 | 58 | 55 | 61 |
| White MN | PFR | Booster | 2081936 | 139 | 13825144 | 6465 | 86 | 83 | 88 |
| White MN | MOD | Complete >26 | 4794767 | 768 | 13825144 | 6465 | 66 | 63 | 68 |
| White MN | MOD | Booster | 612267 | 49 | 13825144 | 6465 | 83 | 77 | 87 |
| Diabetes | PFR | Complete >26 | 538192 | 306 | 1244587 | 1096 | 35 | 27 | 43 |
| Diabetes | PFR | Booster | 290177 | 43 | 1244587 | 1096 | 83 | 77 | 88 |
| Diabetes | MOD | Complete >26 | 534738 | 232 | 1244587 | 1096 | 51 | 43 | 57 |
| Diabetes | MOD | Booster | 83997 | 12 | 1244587 | 1096 | 84 | 71 | 91 |
| Hypertension | PFR | Complete >26 | 1524436 | 595 | 3056716 | 2068 | 42 | 37 | 47 |
| Hypertension | PFR | Booster | 866678 | 92 | 3056716 | 2068 | 84 | 81 | 87 |
| Hypertension | MOD | Complete >26 | 1459165 | 495 | 3056716 | 2068 | 50 | 45 | 55 |
| Hypertension | MOD | Booster | 239004 | 35 | 3056716 | 2068 | 78 | 70 | 85 |
| Heart Disease | PFR | Complete >26 | 512684 | 372 | 1501792 | 793 | -37 | -55 | -21 |
| Heart Disease | PFR | Booster | 332199 | 59 | 1501792 | 793 | 66 | 56 | 74 |
| Heart Disease | MOD | Complete >26 | 528643 | 319 | 1501792 | 793 | -14 | -30 | 0 |
| Heart Disease | MOD | Booster | 91796 | 20 | 1501792 | 793 | 59 | 36 | 74 |
| Cancer | PFR | Complete >26 | 441634 | 170 | 909411 | 399 | 12 | -5 | 27 |
| Cancer | PFR | Booster | 328274 | 46 | 909411 | 399 | 68 | 57 | 76 |
| Cancer | MOD | Complete >26 | 388647 | 117 | 909411 | 399 | 31 | 16 | 44 |
| Cancer | MOD | Booster | 100660 | 24 | 909411 | 399 | 46 | 18 | 64 |
| COPD | PFR | Complete >26 | 372208 | 284 | 1380847 | 1251 | 16 | 4 | 26 |
| COPD | PFR | Booster | 194307 | 42 | 1380847 | 1251 | 76 | 68 | 82 |
| COPD | MOD | Complete >26 | 374250 | 201 | 1380847 | 1251 | 41 | 31 | 49 |
| COPD | MOD | Booster | 58989 | 18 | 1380847 | 1251 | 66 | 46 | 79 |

Note: MOD, Moderna; PFR, Pfizer; VE, vaccine effectiveness. Complete >26 indicates individuals more than 26 weeks since a second dose. Booster indicates inviduals at least two weeks after a booster dose of Pfizer or Moderna.
